# Severity and Correlates of Mental Fog in People with Traumatic Brain Injury

**DOI:** 10.1101/2022.04.04.22273405

**Authors:** Tyler Bell, Michael Crowe, Thomas Novack, Richard D. Davis, Despina Stavrinos

**Author notes:** Correspondence concerning this article should be addressed to Despina Stavrinos, Department of Psychology, University of Alabama at Birmingham, Birmingham, AL 36688. Author Note Tyler Reed Bell, PhD, Department of Psychiatry, University of California, San Diego,; Michael Crowe, PhD, Department of Psychology, University of Alabama at Birmingham,; Thomas Novack, PhD,; Richard Davis, MD., Sports Medicine, University of Alabama at Birmingham,; Despina Stavrinos, PhD, Department of Psychology, University of Alabama at Birmingham.

## Abstract

**Objective:** Alongside objective performance declines, self-reported cognitive symptoms after traumatic brain injury (TBI) abound. Mental fog is one symptom that has been underexplored. The current project investigated mental fog across two studies of individuals with mild traumatic brain injury and moderate-to-severe traumatic brain injury to close our knowledge gap about differences in severity. We then explored the cognitive and affective correlates of mental fog within these groups.

**Methods:** Using between-groups designs, the first study recruited individuals with acute mild TBI (*n* = 15) along with a healthy control group (*n* = 16). Simultaneously, a second study recruited persons with post-acute moderate-to-severe TBI, a stage when self-reports are most reliable (*n* = 15). Measures across the studies were harmonized and involved measuring mental fog (Mental Clutter Scale), objective cognition (Cogstate^®^ and UFOV^®^), and depressive symptoms. In addition to descriptive group difference analyses, nonparametric correlations determined associations between mental fog, objective cognition, and depressive symptoms.

**Results:** Results revealed higher self-reported mental fog in acute mild TBI compared to healthy controls. And though exploratory, post-acute moderate-to-severe TBI also appears characterized by greater mental fog. Correlations showed that mental fog in mild TBI corresponded to greater depressive symptoms (*r* = .66) but was unrelated to objective cognition. By contrast, mental fog in moderate-to-severe TBI corresponded to poorer working memory (*r* = .68) and slowed processing speed (*r* = -.55) but was unrelated to depressive symptoms.

**Conclusion:** As a common symptom in TBI, mental fog distinguishes individuals with acute mild TBI from uninjured peers. Mental fog also appears to reflect challenges in recovery, including depressive symptoms and objective cognitive problems. Screening for mental fog, in addition to other cognitive symptoms, might be worthwhile in these populations.

---

Every year, two million people in the United States suffer external, kinetic force to the brain-altering function and creating pathology known as traumatic brain injury (TBI) (Frost et al., 2013). TBI is typically classified as mild, involving a loss of consciousness less than 30 minutes of lost consciousness and significant post-concussive symptoms. Fewer cases are classified as moderate-to-severe, involving more than 30 minutes of lost consciousness, amnesia, and observable brain damage (Vos et al., 2002). Across severities, physical and emotional issues emerge, but considerable difficulties arise for cognition. Such issues entail trouble remembering or learning information, thinking quickly, paying attention, and everyday problem solving (Schretlen & Shapiro, 2003). Considerable research has described the wide array of objective (task-measured) cognitive difficulties experienced after a TBI. Still, we know less about the full profile of cognitive symptoms experienced, and what they might tell us about the individual injured before in-depth assessment.

Self-reported cognitive symptoms can range across multiple abilities and likely depend on injury severity (Schmand et al., 1996). For mild TBI, common symptoms include troubles remembering, concentrating, and slowed thinking (de Boussard et al., 2005; Ponsford et al., 2011). Moreover, while most objective cognitive problems resolve weeks to months after injury, cognitive symptoms endure up to 8 years after injury and are greater than their peers’ reports (*d* = .75) (Dean et al., 2012). Thus, these symptoms either pick up on suboptimal cognitive performance above impairment or other unresolved difficulties during long-term recovery. Such appears the case also for moderate-to-severe cases, where self-reported symptoms predominantly involve troubles in memory and problem solving (Corrigan et al., 2004). This likely also affects functional outcomes long term as these symptoms last anywhere from two (*d* = .62) to twenty-four years (*d* = .12) after moderate-to-severe TBI (Corrigan et al., 2004; Gardner et al., 2017; Hart et al., 2005).

One cognitive symptom receiving little attention is mental fog. As a common symptom reported within community patient groups and clinics, mental fog involves self-reported problems in memory, attention, and processing speed coupled *with a lack of mental clarity* (Katz et al., 2004; Nelson & Esty, 2015; Theoharides et al., 2015). Evidence for this symptom comes from post-concussion symptom screeners originally asking about feeling “fogginess” or “in a fog,” screenings which showed a ranging prevalence in the acute mild TBI period (17 to 81.2%). Clinicians have labeled mental fog as a significant health challenge in patients with mild TBI and key symptom for diagnosis by the International Conference on Concussion in Sport (McCrory et al., 2009). However, more research is needed to determine if this symptom is more severe in people with mild TBI by inclusion of control group comparison. Also, there is a great lack of understanding how this symptom is manifested in persons with moderate-to-severe TBI, likely due to practical reasons. Studies on the effects of acute TBI on mental fog likely exclude moderate-to-severe cases due to substantial awareness deficits, which would render self-reported symptoms unreliable. Thus, studies should focus on the post-acute phase where reports become more clinically useful and might help understand residing functional issues (Stuss et al., 1999; Sherer et al., 2005) (Nakase-Thompson et al., 2005). Hence, there is a need to detail the severity of mental fog during periods where it would be most advantageous to assess, i.e., acute mild TBI and post-acute moderate-to-severe TBI. In these periods, objective cognitive issues reside, and reliable self-reports appear obtainable.

In addition to better describing the severity of mental fog in persons in these critical periods, there is a need to understand what these symptoms tell us about a person. Does greater mental fog reflect ongoing issues in objective cognition or depressive symptoms? Cognitive symptom literature details two possibilities (Hertzog, Hülür, Gerstorf, & Pearman, 2018): One, individuals may accurately monitor their cognitive problems after injury (*accurate monitoring hypothesis*). If so, symptoms should correlate with objective cognitive measures. Two, cognitive symptoms might otherwise represent an extension of negative self-thinking, such as that induced by depressive symptoms (*constructed judgment hypothesis*). Support for both mechanisms has been shown in the broader literature and appears to depend on the samples studied and cognitive problems queried (Hertzog et al., 2018; Hülür, Willis, Hertzog, Schaie, & Gerstorf, 2018). Thus, associations might differ between individuals with mild TBI and moderate-to-severe TBI.

To close this critical knowledge gap, our investigatory aims were two-fold: One, we sought to use a comprehensive measure of mental fog to explore symptom severities in people with mild and moderate-to-severe TBI. Second, we strived to understand what factors contribute to these reports. Are people accurately monitoring their cognition, or are they constructing negative judgments based on depressive symptoms? Analyses will involve integrating data from two recent studies on mild and moderate-to-severe traumatic brain injury conducted with harmonized measures.

## METHODS

### Participants

The first study occurred from Fall 2016 to Summer 2018, where we recruited 15 individuals ages 16 to 25 with a mild TBI within two weeks of injury from local concussion clinics. Individuals were eligible if they reported substantial symptoms on the Post-Concussive Symptom Scale (individuals scoring ≥ 13) (Lovell et al., 2006) and experienced one major TBI symptom (e.g., loss of consciousness ≤ 30 minutes; loss of balance or motor coordination; disorientation or confusion; loss of memory; or dizziness; Kay et al., 1993). In addition to these participants, the first study recruited 16 healthy controls via community advertisements matching the mild TBI group on age and gender characteristics.

In the second study, we recruited 15 adult participants with a diagnosed moderate-to-severe TBI, ages 21 to 50 years, referred from the UAB Traumatic Brain Injury Model Systems (injury occurred < within the past 24 months). All procedures were ethically approved by our Institutional Review Board. Further details about these studies have been published elsewhere (McManus, Bell, & Stavrinos, 2019; McManus, Cox, Vance, & Stavrinos, 2015; Newton et al., 2018; Stavrinos et al., 2019).

### Measures

Studies collected information on demographics, mental fog, objective cognition, and depressive symptoms via harmonized measures described below:

### Demographics

From telephone screening, participants provided information on their age, gender, race, and ethnicity. Lastly, we collected the date of the most recent TBI.

### Mental fog

Mental fog was measured by the 16-item Mental Clutter Scale (MCS) (Leavitt & Katz, 2011). The MCS was developed to provide a detailed scale of mental fog over two dimensions: self-reported symptoms of general cognitive problems and lack of mental clarity. Questions asked participants to rate how much they have experienced different issues from 1 “Not at all” to 10 “All the time.” Example items for general cognitive symptoms included trouble with “concentration,” “memory,” or “mental speed,” whereas mental clarity included self-reported problems with “spaciness,” “fogginess,” or “information overload.” The research revealed that these two dimensions (8-items each) contained good factor stability (Leavitt & Katz, 2011). However, a one-factor score also produces strong reliability while mirroring criteria for mental fog, i.e., both subjective cognitive problems and a lack of mental clarity (Leavitt & Katz, 2011). Our study demonstrated high reliability for a total score across groups (αs ranged from .95 to .96). This total score ranges from 16 to 160, where higher values indicate greater mental fog.

### Objective cognition

#### Cogstate Brief Battery^®^

An objective evaluation of cognitive performance was obtained by the Cogstate Brief Battery^®^ (Collie et al., 2003). This battery is derived from the general Cogstate Battery^®,^ which is comprehensive and tests several domains (see www.cogstate.com). However, for brevity, the current study used a brief battery, which only takes 10 to 15 minutes to complete. The Cogstate Brief Battery^®^ consisted of four tasks:

1. The Detection task was a simple reaction time task in which participants pressed a “YES” key (Letter K) when they saw a card turned face-up on the screen.
2. The Identification task was a choice-reaction time task in which participants determined if a car was red or black and pressed the appropriate key.
3. The One-Back task is a working memory task like the n-back; in this task, participants selected if the card presented to them was the same as the one just before.
4. Lastly, in the Learning task, participants selected if a card presented was ever presented in the deck before; this required intact memory and learning ability.

Each task had a set of 1 to 3-minute practice trials to ensure comprehension of the task. To ensure optimum performance, participants wore a headset for auditory performance feedback (e.g., which makes a harsh tone for wrong answers and a light sound for correct answers). Scores were calculated using a proprietary algorithm incorporating speed, accuracy, hits, misses, and anticipations. Tests show strong construct validity with other neuropsychological measures (Maruff et al., 2009).

#### Useful Field of View^®^

Useful Field of View (UFOV^®^) (Ball & Owsley, 1993) also captured objective cognition and has been used previously in persons with TBIs (Novack et al., 2006). UFOV^®^ consisted of four tasks capturing processing speed and forms of executive function.

1. UFOV^®^1 – Stimuli Identification: Participants quickly determined if they viewed a “car” or “truck” within milliseconds of exposure. This task captured speed of processing.
2. UFOV^®^2 – Divided Attention: Participants shifted between identifying a car or truck in the center and remembering the location of a car in the periphery. The location of the car in the periphery occurred anywhere on an eight-spoke spiral around the center stimuli. As named, this task estimated divided attention but partly captured set-shifting ability.
3. UFOV^®^3 – Selective Attention I: Participants completed the same task as UFOV^®^2 but in the presence of distracting stimuli (47 triangles) across the screen.
4. UFOV^®^4 – Selective Attention II: The fourth subtest also tested selective attention in the presence of distractors (47 triangles) but with a new task involving the center stimuli. Participants decided if two stimuli in the center were the same (two cars or two trucks shown) or different (car and truck shown) while determining the location of the peripheral car as before. However, it involved the introduction of a novel task that increased difficulty.

Each subtest comprised visual demonstrations and a 2-minute practice to verify task comprehension. During their performance, the software provided an exposure threshold where 75% of responses were correct. These scores approximated optimal ability for each cognitive domain.

### Depressed mood

The Profile of Mood States (POMS) captured depressive symptoms for both injury and healthy control groups. The POMS was a 37-item instrument that allowed participants to denote feelings “since their injury” for the mild and moderate-to-severe TBI groups and “in the last two weeks” for the healthy control group. Participants rated how frequently they experienced various symptoms using the following Likert-type scale: “1-not at all,” “2-a little,” “3-moderately,” “4-quite a bit,” and “5-extremely.” This format provided a quickly answerable instrument with high factorial, face, and construct validity (McNair et al., 1971). To control for between- and within-group differences in negative affect, we used the depression (POMS-Dep) subscale from this instrument. This subscale includes feelings of being unhappy, sad, hopeless, discouraged, miserable, helpless, and worthless. POMS-Dep scores range from 8 (no depressive symptoms) to 40 (severe depressive symptoms) with excellent internal consistency in the current study (αs ranged from .88 to .94).

### Post-Concussive Symptoms

The Post-Concussive Symptom Scale (Lovell et al., 2006) measured injury severity. For 22 listed symptoms, participants rated their occurrence from none (0) to severe (6). Symptoms entailed cognitive (4 items), somatic (14 items), and mental/psychological difficulties (4 items). Summed responses ranged from 0 (no concussion symptoms) to 132 (high concussion symptoms). For healthy controls, scores represent general health problems. Internal consistency was high (αs ranged from .89 to .95).

### Procedure

Participants were assessed within two weeks to confirm mild TBI and within 24 months for moderate-to-severe TBI. After verifying eligibility, we scheduled participants for an appointment in the laboratory. At this session, participants reported their level of mental fog (MCS), depressive symptoms (POMS-DEP), and post-concussive symptoms (PCSS). Afterward, participants completed the Cogstate Brief Battery^®^ and UFOV^®^. We remunerated participants for their time.

### Statistics data analysis

For our first set of analyses, we calculated descriptive statistics on all variables using SPSS version 25 (see Tables 1 to 2). An ANOVA then examined the effect of group (control, mild TBI, and moderate-to-severe TBI) on the continuous measure of mental fog (MCS total). If provided a significant omnibus test, Tukey’s post-hoc tests were then conducted to determine pairwise differences (control versus mild TBI, control versus moderate-to-severe TBI, mild TBI versus moderate-to-severe TBI). Because of the small sample possible non-normality of mental fog, this was followed by a Kruskal– Wallis test, which inspected group differences based on rank. Secondly, because the moderate-to-severe TBI group was older based on original study aims, a sensitivity analysis using a generalized linear model tested if group differences in mental fog remained after accounting for age differences. Comparisons on objective cognition are provided in Table 1. However, they are not discussed directly as they are beyond the scope of this project (although in expected directions).

**Table 2.**
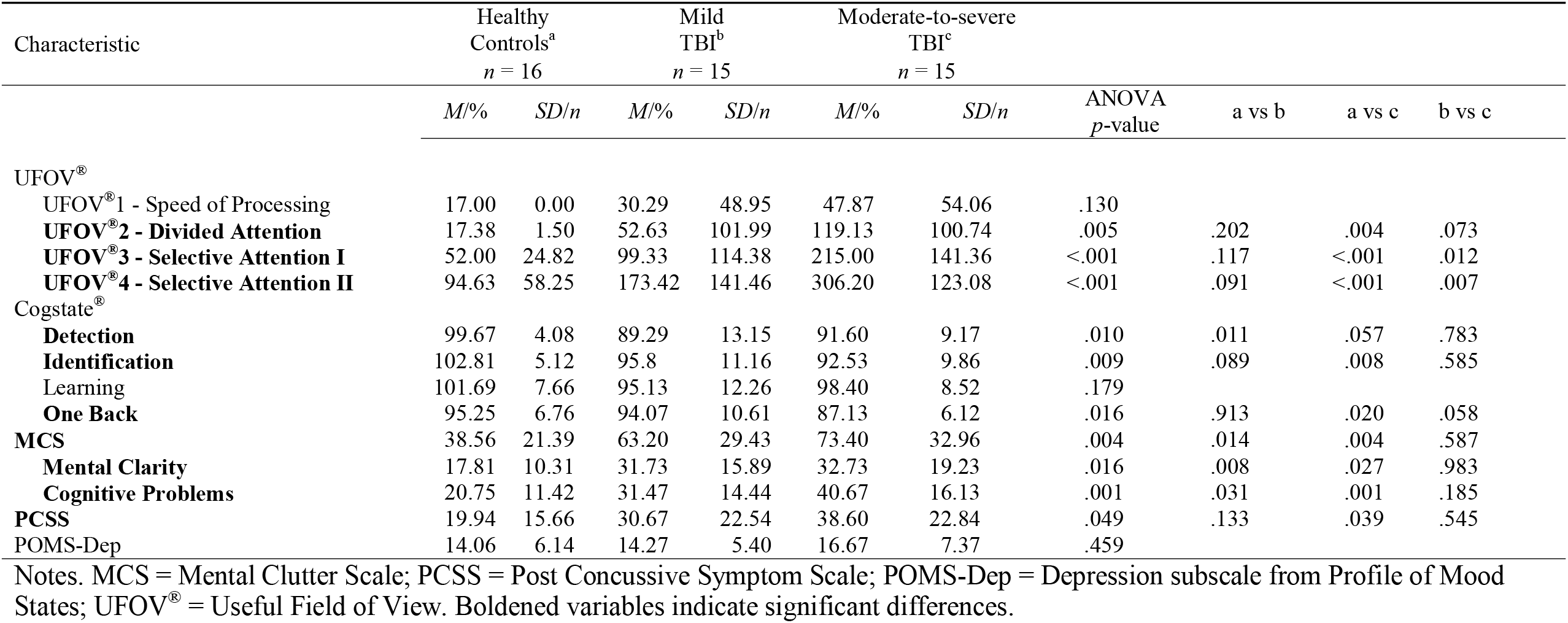
Shared Study Outcomes.

For primary analyses, we tested group-specific relations between mental fog with objective cognition (processing speed time, processing speed accuracy, working memory, and memory) and depressive symptoms using Spearman correlations. Correlations with objective cognition involved an adjustment for depressive symptoms as a second step.We reported *p*-values and effect sizes for all analyses and determined significance at the .05 level.

## Results

### Descriptives

#### Demographics

Participant demographics are shown in Table 1. For the mild TBI group (*n* = 15), the average participant age was 16.73 years (*SD* = 0.80, range: 16 to 19). This group was predominately female (60.0%, *n* = 9) and Caucasian (80.0%, *n* = 12). The healthy control group (*n* = 16) appeared successfully matched to the mild TBI group. Meanwhile, individuals with a moderate-to-severe TBI were predominately young to middle-aged adults (*M*_age_ = 33.19 years, *SD* = 8.74, range = 20 to 50) – significantly older than our mild TBI and control samples (*F*(2,43) = 51.72, *p* < .001, *η*^2^ = .71). Regarding other personal characteristics, the sample consisted of a slight male majority (56.3%, *n* = 9) who were predominantly Caucasian (73.3%, *n* = 11). The moderate-to-severe TBI group were similar on sex (*X*^2^(2) = 1.33, *p* = .514) and race proportions (*X*^2^(2) = .20, *p* = .905) compared to the mild TBI and healthy control groups. Because age was the only significant difference across groups, age was included as a covariate in later our descriptive comparisons.

#### Group differences in mental fog

After conducting a one-way ANOVA, we found a significant effect of group on mental fog (*F*(2,43) = 6.29, *p* = .004; *η* ^2^ = .23; 95%CI[.03 to .40]) (shown in Figure 1 and Table 3). Post-hoc tests confirmed that individuals with mild TBI reported higher mental fog compared to healthy controls (*M*_Diff_ = 24.64, *p* = .014; *d* = .96) as did individuals with moderate-to-severe TBI (*M*_Diff_ = 34.84, *p* = .004, *d* = 1.25). No significant difference emerged between individuals with mild TBI and moderate-to-severe TBI (*M*_Diff_ = 10.20, *p* = .327). Sensitivity analyses showed that these patterns of results survived nonparametric testing using a nonparametric Kruskal-Wallis test as well as adjustment for depressive symptoms and age (*p*s < .01).

**Figure 1.**
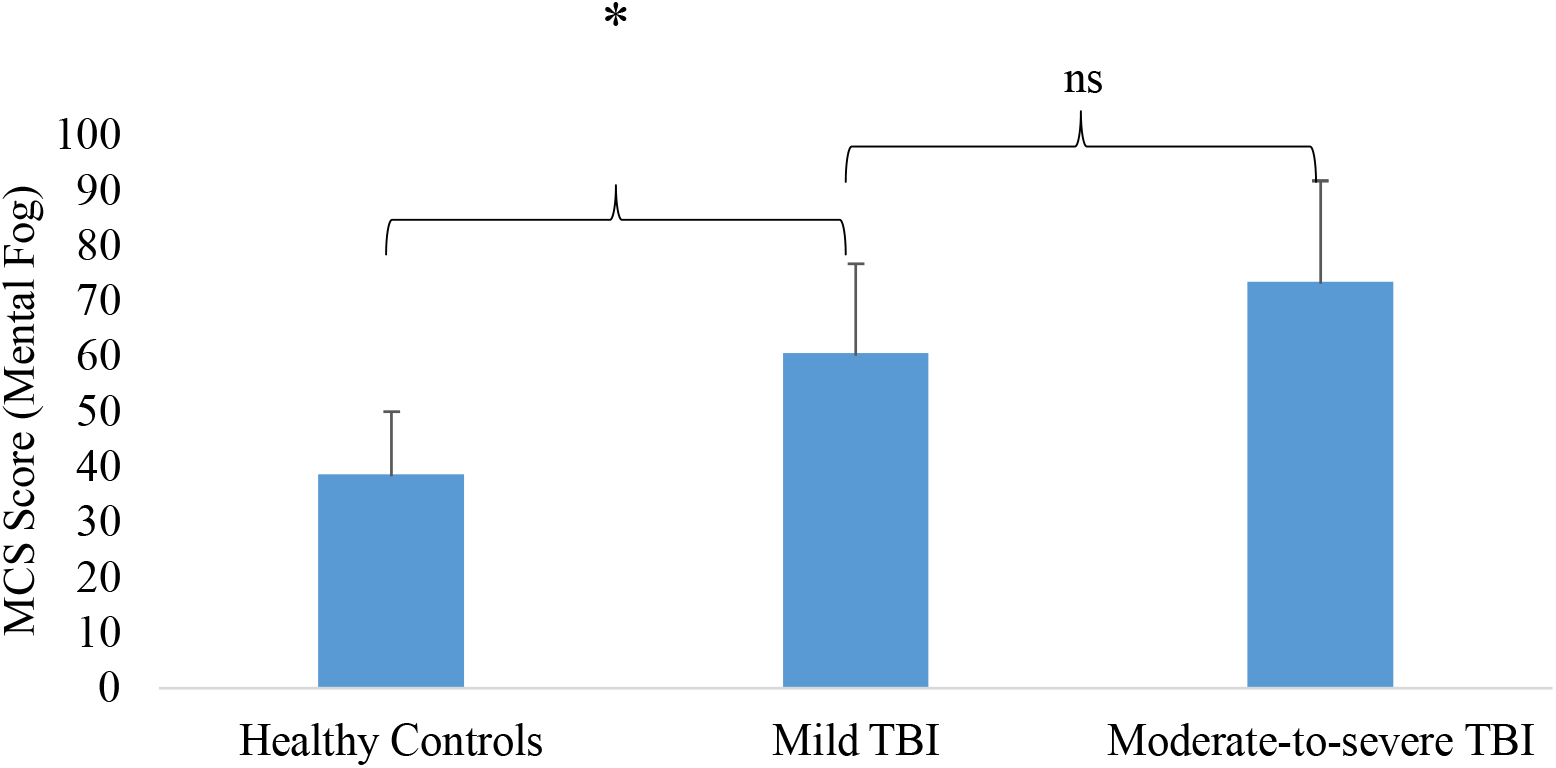
Group differences on mental fog.

**Table 3.** Correlations between Mental Fog and Objective Cognitive Tasks in Healthy Controls (*n* = 16).

|  | 1 | 2 | 3 | 4 | 5 | 6 | 7 | 8 | 9 | 10 | 11 | 12 | 13 |
| --- | --- | --- | --- | --- | --- | --- | --- | --- | --- | --- | --- | --- | --- |
| 1. MCS | 1.00 | .65** | .57* | .11 | -.38 | .10 | .08 | -.15 | -.25 | 0 | .36 | .35 | .34 |
| 2. POMS-Dep |  | 1.00 | .81** | -.04 | -.34 | .24 | .29 | -.13 | -.22 | 0 | -.03 | .15 | .12 |
| 3. PCSS |  |  | 1.00 | -.08 | -.49<sup>†</sup> | .19 | -.01 | -.17 | -.06 | 0 | .08 | .10 | .05 |
| 4. Age |  |  |  | 1.00 | -.23 | .22 | .13 | -.17 | -.13 | 0 | .33 | -.08 | .31 |
| 5. Sex |  |  |  |  | 1.00 | -.10 | -.10 | .07 | -.02 | 0 | -.20 | -.25 | -.42 |
| 6. Detection |  |  |  |  |  | 1.00 | .67* | .33 | .53** | 0 | -.03 | -.22 | -.29 |
| 7. Identification |  |  |  |  |  |  | 1.00 | .44 | .27 | 0 | -.19 | .24 | -.05 |
| 8. Learning |  |  |  |  |  |  |  | 1.00 | .07 | 0 | -.16 | -.25 | -.43 |
| 9. One Back |  |  |  |  |  |  |  |  | 1.00 | 0 | -.19 | -.14 | -.35 |
| 10. UFOV<sup>®</sup>1 |  |  |  |  |  |  |  |  |  | 1.00 | 0 | 0 | 0 |
| 11. UFOV<sup>®</sup>2 |  |  |  |  |  |  |  |  |  |  | 1.00 | .06 | .42 |
| 12. UFOV<sup>®</sup>3 |  |  |  |  |  |  |  |  |  |  |  | 1.00 | .59* |
| 13. UFOV<sup>®</sup>4 |  |  |  |  |  |  |  |  |  |  |  |  | 1.00 |

Notes. All correlations are Spearman rho. MCS = Mental Clutter Scale; PCSS = Post-Concussive Symptom Scale; POMS-Dep = Depression subscale from the Profile of Mood States; UFOV = Useful Field of View. <sup>†</sup> $p < .10$ , \* $p < .05$ , \*\* $p < .01$ , \*\*\* $p < .001$

### Correlates of mental fog

#### Healthy controls

Mental fog did not associate with any scores on UFOV^®^ or Cogstate^®^ in healthy controls (|*r*s_sp_| range: .04 to 42, all *p*s > .10). Although greater mental fog reflected worse divided attention after controlling for depressive symptoms (*r*_sp_ = .57, *p* = .044).

#### Mild TBI

We calculated Spearman rank correlations between mental fog and objective cognition in mild TBI, as shown in Table 4. For individuals with mild TBI, higher mental fog corresponded to slower speed of processing (UFOV^®^1; *r*_sp_ = .63, *p* = .012) but was unrelated to all other measures (|*r*s_sp_| range: .02 to .35, *p*s > .20). After adjusting for depressive symptoms, the relationship between mental fog and speed-of-processing was nonsignificant (*r*_sp_ = .25, *p* = .488).

**Table 4.**
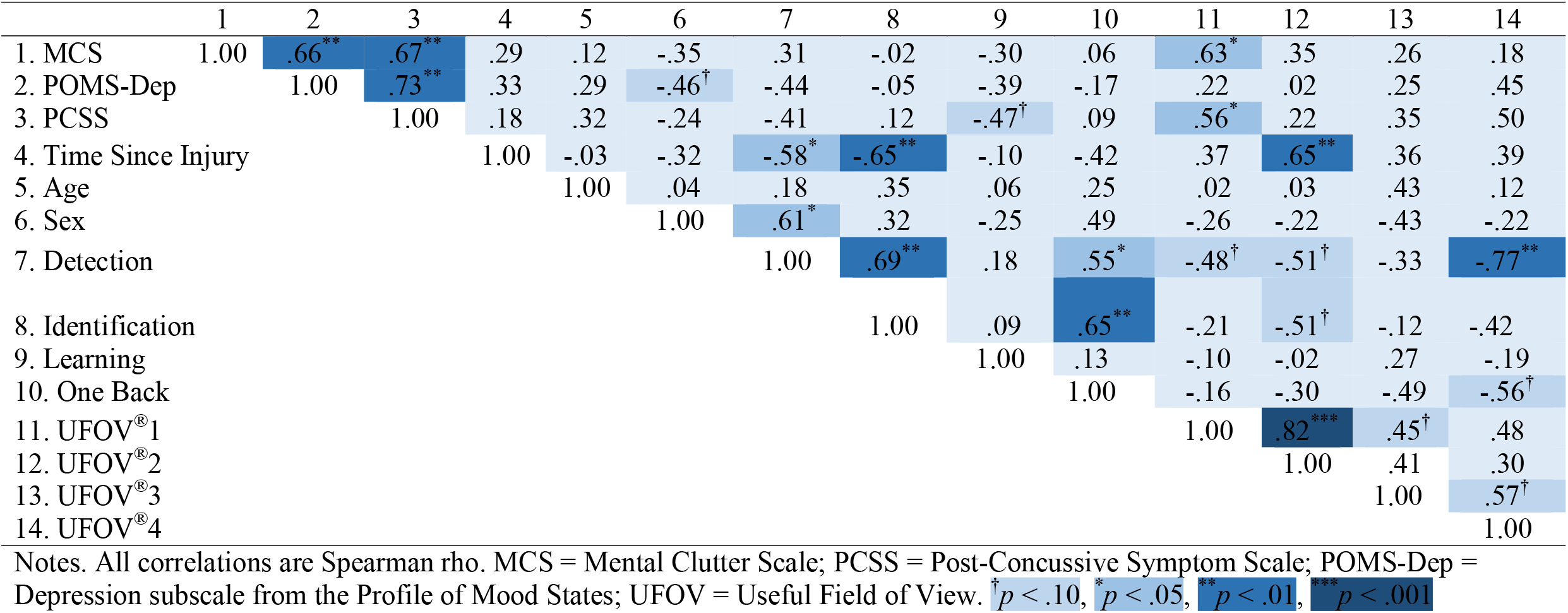
Correlations between Mental Fog and Objective Cognitive Tasks in Persons with Mild TBI (*n* = 15).

#### Moderate-to-severe TBI

We calculated Spearman rank correlations between mental fog and objective cognition in moderate-to-severe TBI, seen in Table 5. Greater mental fog related to slower processing speed indexed by lower scores on the Detection task (*r*_sp_ = -.55, *p* = .033). Also, mental fog also negatively related to memory (*r*_sp_ = -.55, *p* = .034) and working memory (*r*_sp_ = -.68, *p* = .006) indexed by lower scores on the Learning and One Back tasks, respectively (see Figure 2 and 3). Greater mental fog severity marginally associated with slower processing speed indexed by lower scores on the Identification task (*r*_sp_ = -.51, *p* = .053).

**Table 5.** Correlations between Mental Fog and Objective Cognitive Tasks in Persons with Moderate-to-severe TBI (*n* = 15).

|  | 1 | 2 | 3 | 4 | 5 | 6 | 7 | 8 | 9 | 10 | 11 | 12 | 13 | 14 |
| --- | --- | --- | --- | --- | --- | --- | --- | --- | --- | --- | --- | --- | --- | --- |
| 1. MCS | 1.00 | .26 | .07 | -.03 | -.16 | .13 | -.55* | .51<sup>†</sup> | -.55* | -.68** | .15 | .05 | -.13 | -.28 |
| 2. POMS-Dep |  | 1.00 | .50<sup>†</sup> | -.39 | .29 | .02 | -.37 | -.66** | .00 | -.35 | .21 | .15 | .42 | -.01 |
| 3. PCSS |  |  | 1.00 | -.25 | .30 | .09 | -.13 | -.31 | .20 | -.20 | .27 | .09 | .23 | -.12 |
| 4. Time since injury |  |  |  | 1.00 | .18 | .13 | .01 | .32 | -.03 | .10 | -.27 | -.29 | -.32 | -.24 |
| 5. Age |  |  |  |  | 1.00 | .24 | -.12 | .09 | -.02 | .40 | -.13 | -.09 | .13 | .10 |
| 6. Sex |  |  |  |  |  | 1.00 | .30 | -.16 | -.14 | -.16 | .15 | .17 | .06 | .17 |
| 7. Detection |  |  |  |  |  |  | 1.00 | .46<sup>†</sup> | .36 | .08 | -.30 | -.10 | -.18 | .19 |
| 8. Identification |  |  |  |  |  |  |  | 1.00 | .25 | .69** | -.28 | -.24 | -.28 | .05 |
| 9. Learning |  |  |  |  |  |  |  |  | 1.00 | .13 | -.58* | -.39 | -.14 | -.19 |
| 10. One Back |  |  |  |  |  |  |  |  |  | 1.00 | .13 | -.03 | .04 | .29 |
| 11. UFOV<sup>®</sup>1 |  |  |  |  |  |  |  |  |  |  | 1.00 | .80*** | .66** | .62* |
| 12. UFOV<sup>®</sup>2 |  |  |  |  |  |  |  |  |  |  |  | 1.00 | .80*** | .77*** |
| 13. UFOV<sup>®</sup>3 |  |  |  |  |  |  |  |  |  |  |  |  | 1.00 | .68** |
| 14. UFOV<sup>®</sup>4 |  |  |  |  |  |  |  |  |  |  |  |  |  | 1.00 |

Notes. All correlations are Spearman rho. MCS = Mental Clutter Scale; PCSS = Post-Concussive Symptom Scale; POMS-Dep = Depression subscale from the Profile of Mood States; UFOV = Useful Field of View. <sup>†</sup> $p < .10$ , \* $p < .05$ , \*\* $p < .01$ , \*\*\* $p < .001$

**Figure 2.**
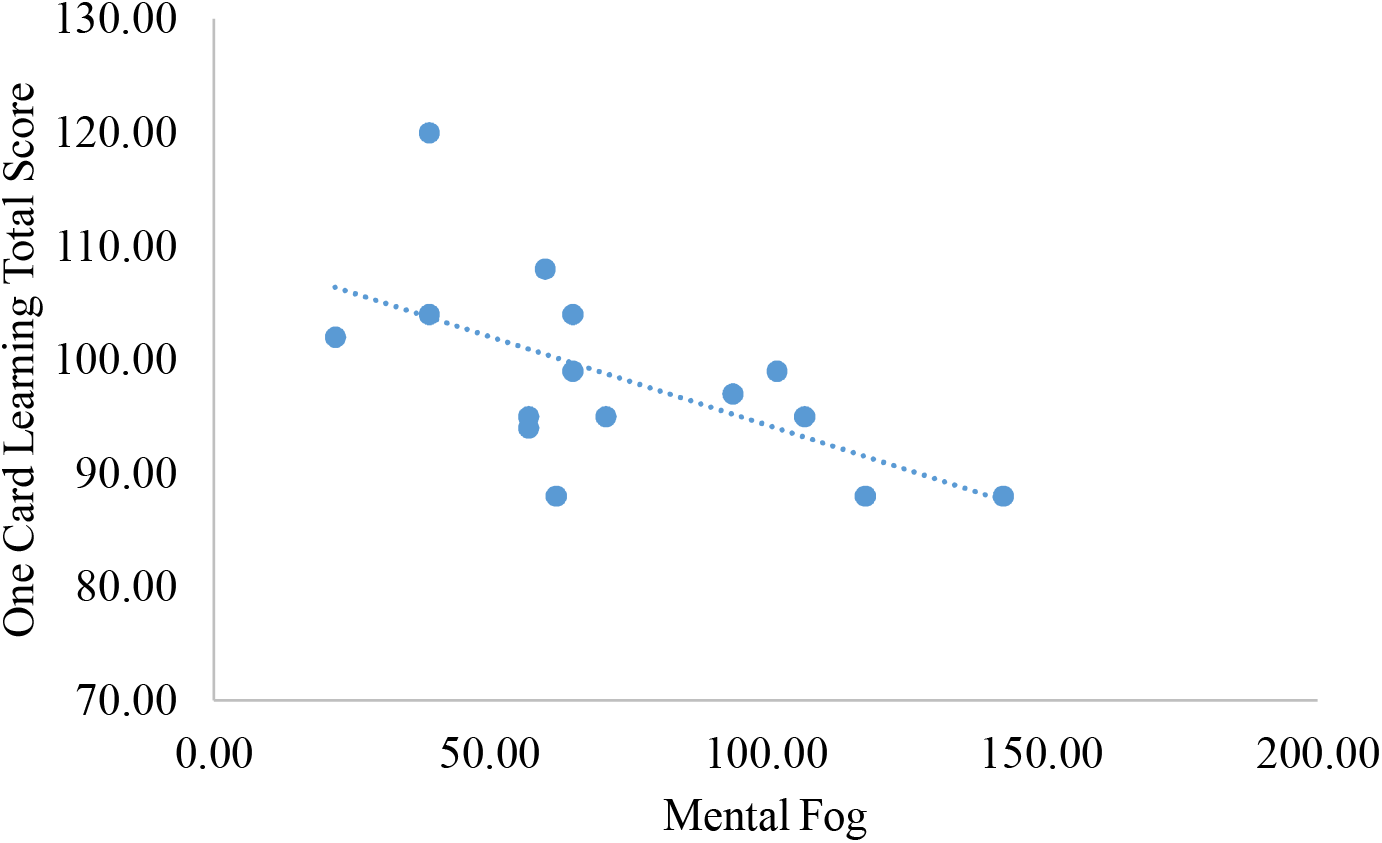
Correlation between Mental Fog and the Cogstate^®^ Learning task.

**Figure 3.**
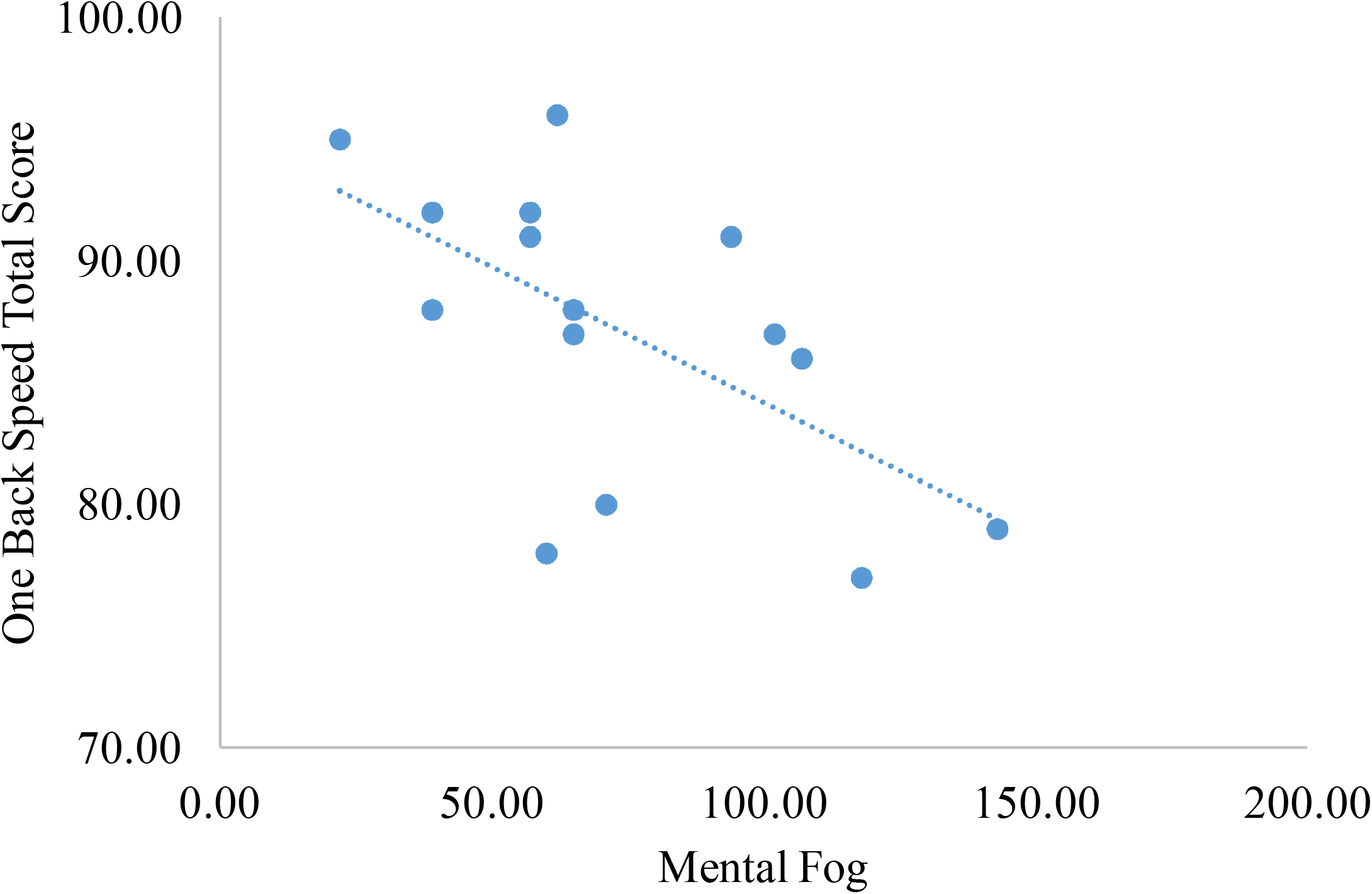
Correlation between Mental Fog and the Cogstate^®^ One Back task.

Next, partial Spearman correlations adjusted for the impact of depressive symptoms. Higher depressive symptoms were unrelated to mental fog (*r*_sp_ = .26, *p* = .349), however. After controlling for depressive symptoms, greater mental fog significantly related to worse scores on the Learning (*r*_sp_ = -.57, *p* = .033) and One Back task (*r*_sp_ = -.65, *p* = .013; see Figure 2 and 3). In addition, mental fog marginally reflected worse processing speed indexed by the Detection (*r*_sp_ = -.51, *p* = .064) and Identification task (*r*_sp_ = -.47, *p* = .094).

## Discussion

Generally, individuals with acute mild TBI reported higher mental fog than healthy controls matched on age and gender proportion (*d* = .96). Findings supplement previous work using single-item assessments (Lovell et al., 2006) and provide novel descriptions of mental fog severity among people with post-acute moderate-to-severe TBI. Overall, results suggest that asking about mental fog might be worthwhile in these groups and perhaps more sensitive than asking about other cognitive symptoms alone. For example, previous work showed a moderate-to-large difference between persons with mild TBI and healthy controls on reported memory loss (*d* = .75; Dean et al., 2012). In comparison, our study found that asking about mental fog provided a more considerable distinction (*d* = .96). This also appeared even more true for persons with a moderate-to-severe TBI (*d* = 1.25). However, further studies will need to test differences in mental fog among people a history of moderate-to-severe TBI within similar age ranges.

As our primary analyses, we explored whether reports of mental fog in each group represented accurate monitoring of cognition or constructed judgments from depressive symptoms. When controlling for depressive symptoms, no association remained between mental fog and objective cognition, supporting the constructed judgment hypothesis in this group. It appears that individuals with mild TBI report higher mental fog due to covarying depressive symptoms rather than coinciding issues in objective cognition. In addition to the constructed judgement hypothesis, it is possible that mental fog and depressive symptoms represent shared symptoms from a common cause. One common cause could be reduced self-efficacy resulting from being unable to keep oneself safe. Another reason could be general worries about recovery and return to normal function extending to symptom reports. Indeed, self-efficacy and general worries show strong links with cognitive symptoms in the broader literature (Aben et al., 2011; Dux et al., 2008). Still, associations remain largely unexplored within traumatic brain injury groups.

Discordance between mental fog and objective cognition in mild TBI contrasts previous studies in this population. As a convergent finding, researchers found that symptoms of inattention and memory loss related more to psychological symptoms than actual memory abilities (Spencer et al. (2010). Conversely, different researchers found that asking about problems in everyday problem-solving (read: executive function) provided ratings more concordant with objectively measured skills such as working memory and set-shifting (Schiehser et al., 2011). Such a divergent finding might reflect differences in how everyday problem-solving is typically assessed compared to most other cognitive symptoms. Specifically, most executive function questions ask participants about disruptions in specific daily behaviors (i.e., *“I have trouble waiting my turn”*) whereas questions on mental fog and memory, for example, rely on global ratings of ability (“Do you feel ‘in a fog’?”; *“Do you have trouble in memory?”*). Behavioral specification might enable participants to disconnect their ratings from self-perceptions towards real-world activities, reducing the influence of psychological symptoms. Future work will be needed to understand if improved item-design could benefit symptom assessments of mental fog, memory, and attention in mild TBI. However, it is also possible that problems in executive functioning are more noticed in people with mild TBI than these other issues. Regardless, mental fog might represent an essential dimension of psychological disruption after mild TBI that uniquely impacts quality of life and everyday function.

Alongside being the first to describe the severity of mental fog in moderate-to-severe TBI, our study simultaneously explored sources for these symptoms. Contrasting the mild TBI group, reports of mental fog aligned more with the accurate monitoring hypothesis in persons with moderate-to-severe TBI. Post-acutely, greater reported mental fog corresponded to lower memory and working memory even after adjusting for depressive symptoms. Although few studies exist in this severity group, this does align with one previous research showing that mental fatigue coincided slower processing speed and reduced working memory (Johansson et al. 2009). Thus, despite lowered awareness in the acute phase, rating of mental fog in the post-acute phase might differentiate persons with moderate-to-severe TBI with residing issues in memory and executive function.

## Limitations

This study was not without limitations. Although the mild TBI group and healthy controls were comparable on age, the moderate-to-severe TBI cohort was much older. Therefore, descriptive comparisons involving this group should be considered with some caution. Fortunately, considerable research shows that age does not significantly affect reports of cognitive symptoms prior to late older age (Devolder & Pressley, 1991), minimizing this concern. Secondly, highly symptomatic mild TBI cases were recruited (> 13 on the PCSS) based on accepted criteria of mild TBI. Nonetheless, high symptomology might influence reporting on other symptom scales like mental fog (Lange et al., 2010). However, depressive symptoms between people with and without mild TBI, reducing this concern. Lastly, sample sizes were modest but bolstered by the well-designed methodology to include representative clinical cases when self-reported mental fog might be most valuable. Moreover, nonparametric statistical techniques were employed to provide conservative testing while providing informative findings for the field.

## Conclusions

After a TBI, individuals experience a plethora of cognitive symptoms, many of which outlast objective cognitive impairments, thus, explaining residual functional concerns. Mental fog is one commonly reported symptom that has been overlooked despite its consideration as a key symptom of mild TBI. Here we validate the ability of mental fog to differentiate people with acute mild TBI from their peers, validating this role as an essential diagnostic symptom. Moreover, we showed that this appears to represent an aspect of disrupted psychological function that might benefit from remediation. Meanwhile, in the post-acute moderate-to-severe TBI group, mental fog represented enduring problems in memory and executive function during the post-acute period rather than depressive symptoms. Future research should consider how mental fog can help explain the longitudinal trajectories of recovery in persons with traumatic brain injury.

## Data Availability

Data is available upon request through a data usage agreement.

## Acknowledgements

The authors would like to thank participants and their caregivers for their time. Additional thanks to JaVarus Humphries and Cierra Hopkins for assistance in data collection.

## Ethical Approval

All procedures performed in studies involving human participants were in accordance with the ethical standards of the institutional and/or national research committee and with the 1964 Helsinki declaration and its later amendments or comparable ethical standards. Informed consent was obtained by all participants.

## Data Availability Statement

The datasets generated during and/or analyzed during the current study are available from the corresponding author on reasonable request.

